# Cellular composition of human early and advanced coronary atherosclerotic lesions

**DOI:** 10.1101/2021.10.07.21264647

**Authors:** Diana Sharysh, Anton V. Markov, Evgeniya S. Grigoryeva, Olga E. Savelieva, Aleksei A. Sleptcov, Aleksei A. Zarubin, Maria S. Nazarenko

**Author notes:** Corresponding author: Maria Nazarenko, Ushayka Embankment 10, Tomsk, 634050, Russia.

## Abstract

**Objective:** Here, we identify and quantify leukocytes, macrophages, endothelial cells (ECs), and vascular smooth muscle cells (VSMCs) with contractile and macrophage-like phenotypes by flow cytometry to compare human early and advanced coronary atherosclerotic plaques.

**Approach and Results:** Sixteen coronary atherosclerotic lesions of 6 patients (3 women, 3 men, age 82 ± 9 years), including one case of restenosis after coronary stenting, were collected at autopsy. The cause of death of all patients was acute myocardial infarction. The lesions were categorized into early (EALs, n = 5) and advanced (AALs, n = 11) atherosclerotic stages. We analyzed a cell suspension stained with antibodies against CD45, CD68, CD31, and αSMA (ACTA2). We noted a decrease in the number of CD45^+^ cells and an increase in the CD45^+^CD68^+^ subpopulation of leukocytes from EALs to AALs. Numbers of CD45^−^αSMA^+^CD68^+^ cells positively correlated with the CD45^+^CD68^+^ macrophage number (r = 0.81; ρ = 0.64; p < 0.05) and the histological type of an atherosclerotic lesion (r = 0.81; ρ = 0.87; p < 0.05). As an interesting case, we analyzed cellular composition of the stented coronary artery and revealed significantly greater numbers of macrophages, αSMA^+^CD68^+^ VSMCs, and ECs in comparison with nonstented plaques.

**Conclusions:** Human early and advanced coronary atherosclerotic lesions differ in their counts of leukocytes and leukocyte subpopulations. For the first time, αSMA^+^CD68^+^ VSMCs were identified in early atherosclerotic stages of coronary arteries. Additionally, the restenotic coronary lesion contains mostly cells and is enriched in ECs, macrophages, and αSMA^+^CD68^+^ VSMCs in particular.

## INTRODUCTION

Atherosclerosis is an age-associated chronic inflammatory disease that involves artery remodeling, significantly reduces quality of life, and leads to high rates of mortality and disability worldwide.^1^ Cellular composition of atherosclerotic arteries was initially investigated by histological techniques with various nonspecific staining methods (e.g., hematoxylin and eosin and Van Gieson’s staining). These histological findings have been the basis for the histological classification of atherosclerotic stages incorporated into clinical practice.^2^

Nonetheless, the most important goal of atherosclerosis research is to predict adverse clinical outcomes of atherosclerotic disease. R. Virmani’s group has suggested a concept of rupture-prone or vulnerable and stable atherosclerotic plaques according to plaque histological characteristics and cellular composition, respectively.^3^ They have stated that the inflammatory component plays a key role in the destabilization of an atherosclerotic plaque and in the development of acute cardiovascular events. On the other hand, vascular smooth muscle cells (VSMCs) form a fibrotic cap and play a protective part.^4^ Therefore, the varying cellular composition of the artery wall determines the clinical impact of atherosclerotic lesions.

Changes in the cellular composition during atherogenesis (from early stages to a complicated plaque) are still unclear. A healthy arterial wall consists of endothelial cells (ECs), VSMCs, fibroblasts, resident immune cells (e.g., dendritic cells [DCs], macrophages, and T and B cells).^5^ Several studies have clarified cell diversity of the healthy arterial wall and identified multiple cellular subpopulations, e.g., pro- and antiatherogenic ECs,^6^ phenotypically modulated Sca1^+^ VSMCs,^7^ and various resident immune cell types.^8^

Atherosclerosis is associated with much higher cellular diversity in the artery wall owing to cell proliferation, phenotype switching, and the recruitment of immune cells in response to low-density lipoprotein cholesterol accumulation and oxidation.^9,10^ Immune cells (including macrophages) perform a pivotal function in the progression and destabilization of human atherosclerotic lesions.^8^ Plaque outcomes are determined by the balance of pro- and anti-inflammatory stimuli produced by these cells.

Recent studies uncovered a significant contribution of VSMCs to plaque formation, progression, and vulnerability.^11^ Nevertheless, due to phenotypic plasticity, VSMCs can play either an adverse or beneficial role in the atherosclerotic plaque.^12^ Different phenotypes of VSMCs (normally contractile, macrophage-like, osteogenic, or synthetic cells or fibromyocytes) are involved in atherosclerosis,^11,13–15^ but VSMC phenotypic switching is poorly studied in humans.

The current knowledge about the pathogenesis of atherosclerosis has been acquired mainly in studies on animal models or *ex vivo* human cell cultures.^7,16–19^ Atherosclerosis is a long-lasting disease, and therefore animal models reproduce the complex human biology insufficiently.^20,21^ There are fundamental differences between the human atherosclerotic disease and mouse models in T- and B-cell composition of atherosclerotic plaques.^22^ Murine atherosclerosis research involving single-cell RNA sequencing (scRNA-seq) technology has revealed high diversity of VSMC phenotypes, e.g., macrophage-like and stemlike. In contrast, scRNA-seq studies on human atherosclerotic plaques have not detected such diversity, except for synthetic and osteochondrogenic phenotypes.^7,11,23,24^

Quantitative and qualitative analyses of cells in the human atherosclerotic plaque to date have been based on a combination of flow cytometry or cytometry by time of flight (CyTOF) and scRNA-seq.^11,23–27^ These reports are cross-sectional (focusing on a single time point) and deal with either carotid or coronary arteries (mostly with advanced plaques). There are several flow-cytometric studies on human carotid, femoral, and aortic atherosclerotic plaques with an analysis of only a few or even a single cell type: VSMCs and immune cells,^28^ immune cells,^26,29^ T cells,^30^ DCs,^31^ mast cells,^32^ and natural killer cells.^33^

There is only scarce information about the complex changes of ECs, leukocytes, macrophages, and VSMC phenotypes between early and advanced stages of coronary atherosclerotic lesions in humans. This work was aimed at identifying these cells in human coronary atherosclerosis and at assessing the differences in cell counts between early and advanced atherosclerotic lesions by flow cytometry. For cell staining, we used a set of markers whose combination allows to distinguish the following cell types: ECs (CD45^−^αSMA^−^CD31^+^), leukocytes (CD45^+^) and separately macrophages (CD45^+^CD68^+^) and contractile (CD45^−^αSMA^+^CD68^−^) and macrophage-like (CD45^−^αSMA^+^CD68^+^) VSMCs.

## MATERIALS AND METHODS

The data that support the findings of this study are available from the corresponding author upon reasonable request.

### Donor material

Sixteen samples of the coronary artery were obtained from 6 donors (3 males, 3 females, age 82 ± 9 years [mean ± SD]) during autopsy. All donors had a cardiovascular disease(s) in their medical history. In all cases, acute coronary syndrome (myocardial infarction) with cardiogenic shock was the cause of death (Table 1).

**Table 1.**
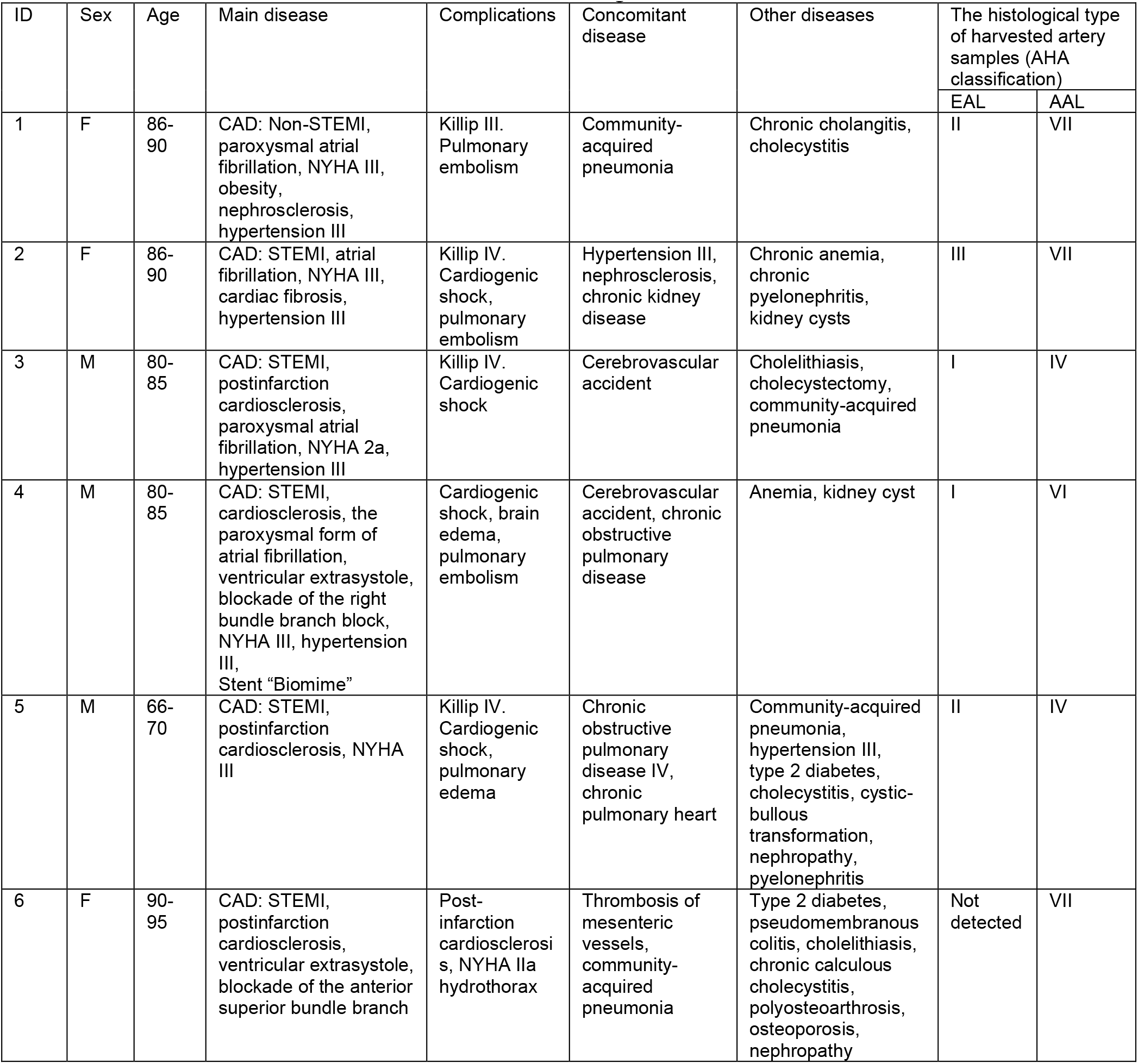
Clinical characteristics of the donors of biological material

| ID | Sex | Age | Main disease | Complications | Concomitant disease | Other diseases | The histological type of harvested artery samples (AHA classification) |  |
| --- | --- | --- | --- | --- | --- | --- | --- | --- |
|  |  |  |  |  |  |  | EAL | AAL |
| 1 | F | 86-90 | CAD: Non-STEMI, paroxysmal atrial fibrillation, NYHA III, obesity, nephrosclerosis, hypertension III | Killip III. Pulmonary embolism | Community-acquired pneumonia | Chronic cholangitis, cholecystitis | II | VII |
| 2 | F | 86-90 | CAD: STEMI, atrial fibrillation, NYHA III, cardiac fibrosis, hypertension III | Killip IV. Cardiogenic shock, pulmonary embolism | Hypertension III, nephrosclerosis, chronic kidney disease | Chronic anemia, chronic pyelonephritis, kidney cysts | III | VII |
| 3 | M | 80-85 | CAD: STEMI, postinfarction cardiosclerosis, paroxysmal atrial fibrillation, NYHA 2a, hypertension III | Killip IV. Cardiogenic shock | Cerebrovascular accident | Cholelithiasis, cholecystectomy, community-acquired pneumonia | I | IV |
| 4 | M | 80-85 | CAD: STEMI, cardiosclerosis, the paroxysmal form of atrial fibrillation, ventricular extrasystole, blockade of the right bundle branch block, NYHA III, hypertension III, Stent "Biomime" | Cardiogenic shock, brain edema, pulmonary embolism | Cerebrovascular accident, chronic obstructive pulmonary disease | Anemia, kidney cyst | I | VI |
| 5 | M | 66-70 | CAD: STEMI, postinfarction cardiosclerosis, NYHA III | Killip IV. Cardiogenic shock, pulmonary edema | Chronic obstructive pulmonary disease IV, chronic pulmonary heart | Community-acquired pneumonia, hypertension III, type 2 diabetes, cholecystitis, cystic-bullos transformation, nephropathy, pyelonephritis | II | IV |
| 6 | F | 90-95 | CAD: STEMI, postinfarction cardiosclerosis, ventricular extrasystole, blockade of the anterior superior bundle branch | Post-infarction cardiosclerosis, NYHA IIa hydrothorax | Thrombosis of mesenteric vessels, community-acquired pneumonia | Type 2 diabetes, pseudomembranous colitis, cholelithiasis, chronic calculous cholecystitis, polyosteoarthritis, osteoporosis, nephropathy | Not detected | VII |

The study protocol was approved by the Local Biomedical Ethics Committee of the Research Institute of Medical Genetics, Tomsk National Research Medical Center (protocol No. 158). The study complied with the Declaration of Helsinki and was conducted in accordance with Russian laws and regulations and institutional policies.

### Examination of human coronary atherosclerotic lesions

Left and right coronary arteries from all donors (total length ∼10–12 cm) were examined with the naked eye and classified as either an early atherosclerotic lesion (EAL; from initial changes up to fatty streaks, without a lipid core) or an advanced atherosclerotic lesion (AAL; a plaque with a lipid core, calcinates, and thrombus). Every sample of the coronary artery was divided into 2 equal pieces: for histological examination and flow-cytometric analysis. The interval between the time of death and sample processing was less than 6 hours.

The tissue samples for histological examination were fixed in 10% formalin, sliced on a microtome, stained with hematoxylin and eosin, and then evaluated by a histopathologist according to American Heart Association (AHA) criteria 2. Consistency between the visual EAL/AAL classification and the histological type of atherosclerosis is demonstrated in Table 1.

The remaining pieces of the artery samples were placed into DMEM (PanEco, cat. No. C410p) and quickly delivered (on ice) to the laboratory for further preparation. The artery samples were cleaned on ice from perivascular fat and soft tissue. The processed samples were placed in a cryopreservation medium (10% dimethyl sulfoxide, 20% fetal bovine serum in DMEM) and cooled slowly down to −80°C in a Mr. Frosty Freezing Container (Thermo Fisher Scientific).

### Disaggregation of human coronary atherosclerotic lesions

Tissue samples were thawed quickly at 37°C, weighed, washed with PBS, and finely minced in 1.5 mL tubes containing 0.5 mL of PBS supplemented with 20 mM HEPES, pH 7.0–7.6. The crude suspension was incubated for 1 hour at 37°C with a digestion mix: 250 U/mL collagenase type IV (PanEco, cat. No. П011-4), 60 U/mL DNase type I (Prospecbio, cat. No. ENZ-417), 60 U/mL hyaluronidase I-S (Sigma-Aldrich, cat. No. H3506), and 100 µg of CaCl_2_ in 2.5 mL of PBS with 20 mM HEPES, pH 7.0–7.6 (Sigma-Aldrich, cat. No. H3375). Next, filtration through a 70 µm BD Falcon™ Cell Strainer (Becton Dickinson, cat. No. 352350), centrifugation (300 × *g* at 4°C for 15 minutes), and resuspension of the cell pellet in 2.5 mL of PBS were performed followed by the second digestion step (200 U/mL collagenase type IV for 20 minutes). After the second filtration and centrifugation step, each cell pellet was resuspended in 1 mL of PBS supplemented with 5% of fetal bovine serum (Biosera, cat. No. FB-1001/500) (FACS buffer) for the blocking of Fc fragments. The numbers and viability of the cells in each suspension were estimated by trypan blue staining in a LUNA II cell counter (Logos Biosystems).

### Flow cytometry of cell suspensions

We determined cell types by flow cytometry on a MoFlo XDP instrument (Beckman Coulter) equipped with 488 nm and 640 nm laser lines. The gating strategy used to identify cell types is outlined in Fig. 1a.

**Figure 1.**
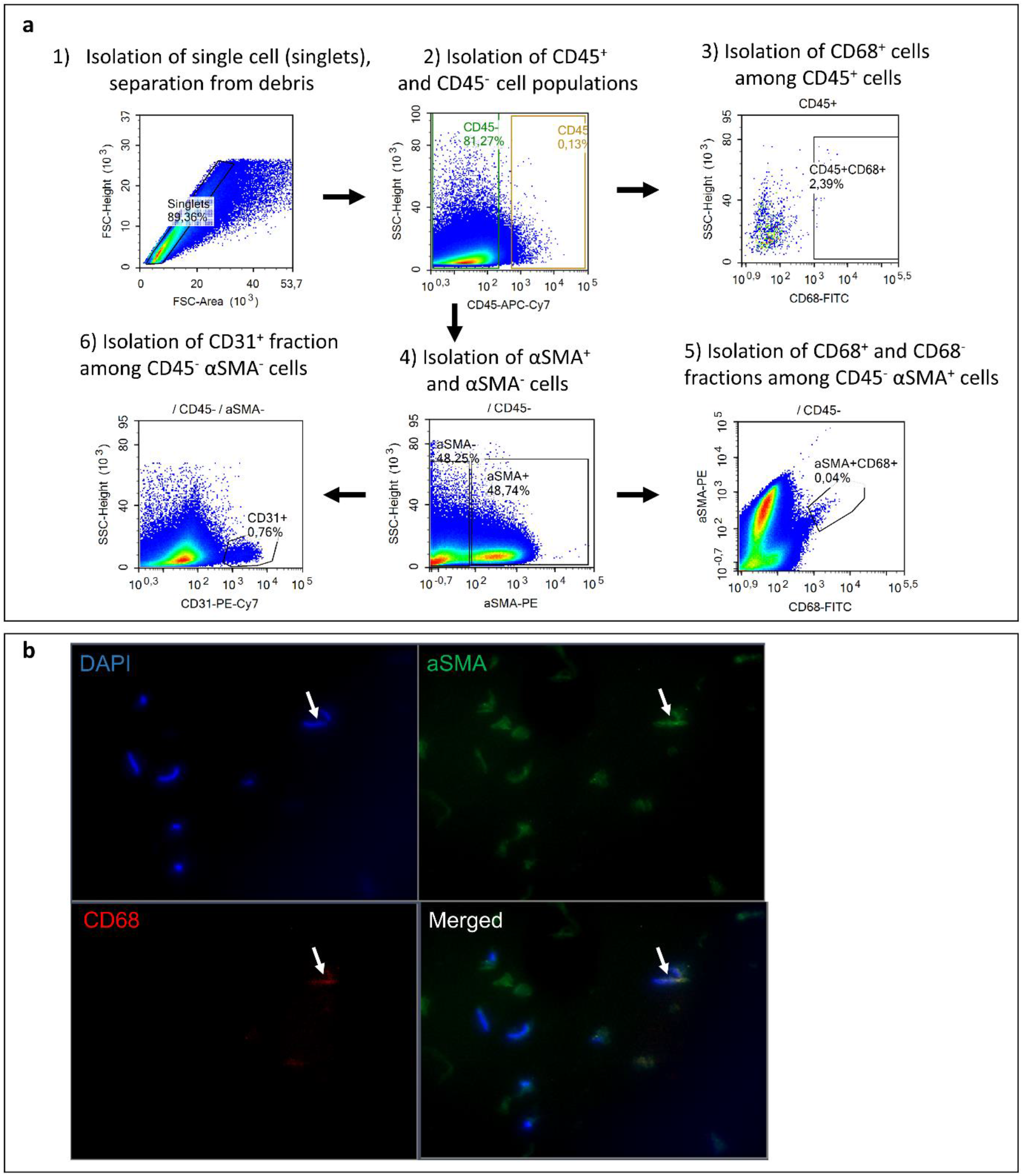
**a**. An outline of the flow-cytometric analysis. The main steps of the analysis include: (1) isolation of singlet cells from debris; (2) isolation CD45^+^ cells from singlets; (3) isolation of CD68^+^ cells from CD45^+^ cells; (4) isolation of αSMA^+^ and αSMA^−^ cells from CD45^−^ cells; (5) isolation of CD68^+^ and CD68^−^ cells from αSMA^+^ cells; (6) isolation of CD31^+^ cells from αSMA^−^ cells. **b**. Fluorescence microscopy of sorted αSMA^+^ cells to validate the flow-cytometric strategy.

The staining of surface and intracellular antigens was performed as follows: 1 mL of a cell suspension was centrifuged at 300 × *g* for 15 minutes. The cell pellet was resuspended in 100 µL of FACS buffer with subsequent addition of monoclonal antibodies against CD45 (antibody conjugated with allophycocyanin H7; Mouse IgG1, κappa, Clone 2D1) and against CD31 (antibody conjugated with phycoerythrin and cyanine 7; Mouse IgG1, κappa, Clone WM59 [RUO], Becton Dickinson, cat. No. 560274 and 563651, respectively) diluted at their optimal titer (please see the Major Resources Table in Supplemental Materials). The suspension was incubated in the dark for 20 minutes.

Next, the cells were permeabilized by means of BD Cytofix/Cytoperm Fixation and Permeabilization Solution (Becton Dickinson, cat. No. 554714) according to the manufacturer’s protocol. Briefly, 1 mL of 1× BD Perm/Wash buffer was added to the cell suspension, and the mixture was centrifuged at 300 × *g* for 10 minutes. The BD Fix/Perm solution was added to the cell pellet followed by vortexing and incubation of the mixture in the dark for 20 minutes at 4°C. The cells were washed with BD Perm/Wash buffer as described above. The cell pellet was resuspended in 50 μL of BD Perm/Wash buffer, then monoclonal antibodies against intracellular antigens αSMA (antibody conjugated with phycoerythrin; Monoclonal Mouse IgG2A Clone 1A4, R&D Systems, cat. No. IC1420P) and CD68 (antibody conjugated with fluorescein isothiocyanate; Mouse BALB/c IgG2b κappa, Becton Dickinson, cat. No. 562117) were added at an optimal titer, and the mixture was incubated in darkness for 15 minutes at 4°C. Finally, the cell suspension was washed with BD Perm/Wash buffer and resuspended in 500 μL of the IsoFlow Sheath Fluid (Beckman Coulter).

The corresponding unstained samples served as a negative control. Compensation was performed using the VersaComp Antibody Capture Bead Kit (cat. No. B22804, Beckman Coulter). In preparation for the downstream data analysis, debris and dead cells were removed. Doublets were identified by means of a ratio of forward scatter signal area (FSC-A) to height (FSC-H) and excluded, followed by gating of cells on the basis of forward and side scatter characteristics.

To identify cellular subsets of interest in the cell suspension samples, a gating strategy based on marker expression patterns was implemented in Summit v5.2 (Beckman Coulter) and NovoExpress v1.4.1 (Agilent Technologies Inc.) software. CD45^+^ cells were designated as leukocytes, CD45^+^CD68^+^ cells as macrophages, CD45^−^αSMA^−^CD31^+^ cells as ECs, and CD45^−^αSMA^+^CD68^−^ and CD45^−^αSMA^+^CD68^+^ cells were assumed to be contractile and macrophage-like VSMCs, respectively. All numbers of cells correspond to singlets.

To check the specificity of our cell sorting, we placed sorted cells on a slide and examined them by fluorescence microscopy (Fig. 1b).

### Statistical analysis

We compared proportions of the identified cell types between EAL and AAL samples by the Mann–Whitney test. The sample of the stented artery was compared with nonstented samples via the one-sample Wilcoxon signed-rank test. Statistical analysis was performed in JASP (v0.11.1, JASP Team). Correlations were evaluated by Spearman’s analysis and Pearson’s linear correlation analysis. Statistical significance was defined as p < 0.05. Variance in the analyzed groups is presented as either the mean ± SD or median (Me) and first (Q1) and third (Q3) quartiles in the form of “Me (Q1; Q3).”

## RESULTS

### The cell types identified in EALs and AALs

The weights of EAL and AAL samples were 0.42 ± 0.18 and 0.64 ± 0.35 g, respectively. EAL samples contained 834000 (482000; 1186000) cells per 0.1 g of wet tissue, with 64 (60; 68)% viability in suspension after disaggregation. AALs consisted of 312000 (52300; 470000) cells per 0.1 g with 55 (49; 63)% viability.

EALs contained CD45^+^ leukocytes at 15.7 (11.9; 16.1)% of all cells, while the proportion of CD45^+^CD68^+^ macrophages was 0.39 (0.28; 0.55)% (Table 2). Resident cells in EALs were as follows: CD45^−^αSMA^+^ VSMCs at 17.6 (10; 41.5)% and CD45^−^CD31^+^ ECs at 0.4 (0.3; 1.4)%. Contractile VSMCs were abundant, 15.9 (7.3; 39.7)%, in comparison with macrophage-like VSMCs, 1.9 (0.8; 2.9)%.

**Table 2.**
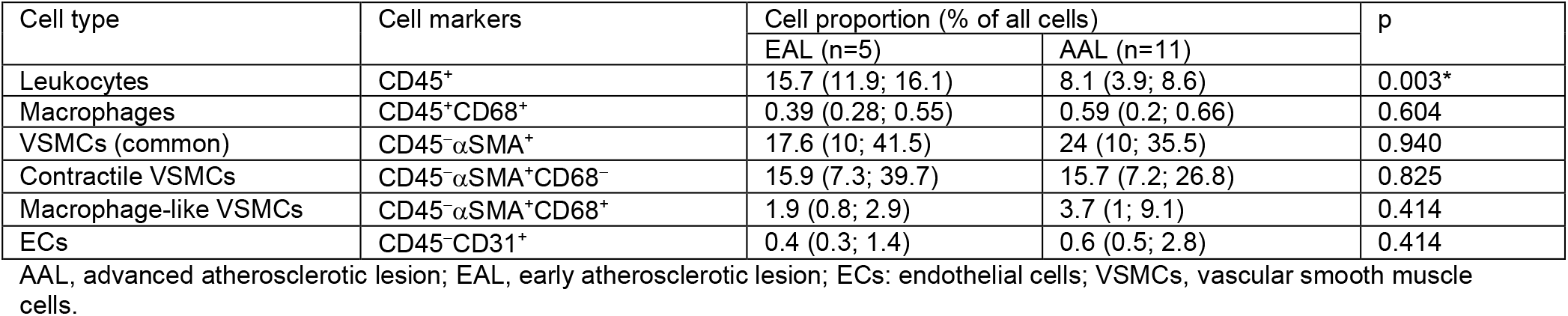
Proportions of cell types identified in early and advanced coronary atherosclerotic lesions

In AALs, we found CD45^+^ leukocytes at 8.1 (3.9; 8.6)%, which included CD45^+^CD68^+^ macrophages at 0.59 (0.2; 0.66)% (Table 2). All CD45^−^αSMA^+^ VSMCs constituted 24 (10; 35.5)% of all cells; 3.7 (1; 9.1)% of VSMCs had the macrophage-like phenotype (CD68^+^), and 15.7 (7.2; 26.8)% of VSMCs had the contractile (CD68^−^) phenotype. The proportion of ECs was 0.6 (0.5; 2.8)% among all cells.

### Changes of immune cells between the EAL and AAL stages

We found a statistically significantly (p = 0.003) greater relative number of CD45^+^ leukocytes in EALs [15.7 (11.9; 16.1)%] than in AALs [8.1 (3.9; 8.5)%; Table 2 and Fig. 2a].

**Figure 2.**
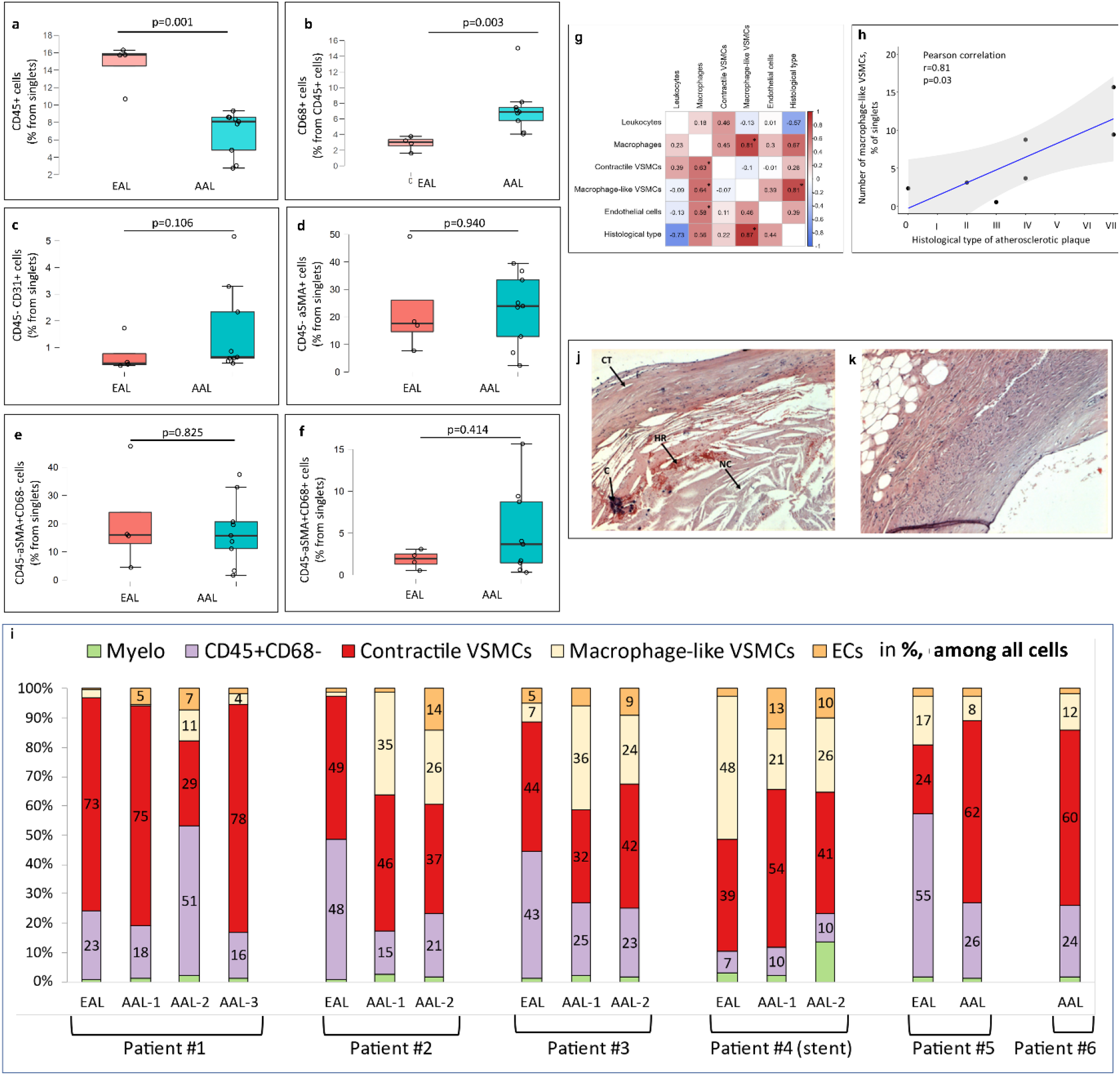
Relative numbers of identified leukocytes in coronary atherosclerotic lesions. **a**. Proportion of all CD45^+^ leukocytes among singlets. **b**. Proportion of CD68^+^ macrophages among CD45^+^ leukocytes. EAL, early atherosclerotic lesion; AAL, advanced atherosclerotic lesion. **c–f**. Proportions of nonimmune cell types identified in early and advanced coronary atherosclerotic lesions. **c**. Endothelial cells. **d**. Vascular smooth muscle cells (common type). **e**. Vascular smooth muscle cells with the contractile phenotype. **f**. Vascular smooth muscle cells with the macrophage-like phenotype. **g**. Heatmaps of correlation between numbers of various cell types and the histological type of atherosclerotic lesions. Top right: Pearson’s linear correlation coefficient. Left bottom: Spearman’s correlation coefficient. An asterisk denotes coefficients with a statistically significant difference from zero: *p < 0.05. **h**. Correlation between the number of macrophage-like VSMCs and the histological type of atherosclerotic lesions (according to AHA classification). **i**. Proportions of cell types identified in different coronary atherosclerotic lesions from the same patient (several patients). Myelo: CD68^+^ macrophages, CD45^+^CD68^−^: non-CD68 leukocytes, contractile VSMCs: αSMA^+^CD68^−^ contractile vascular smooth muscle cells, macrophage-like VSMCs: αSMA^+^CD68^+^ macrophage-like vascular smooth muscle cells, ECs: CD31^+^ endothelial cells; EAL: early atherosclerotic lesion, AAL: advanced atherosclerotic lesion; AAL-(N): ID number of an advanced atherosclerotic plaque, among several from the same patient (where possible). **j, k**. Histological examination of a coronary sirolimus-eluting stent that had been implanted 10 months before the autopsy. **j**. Neoatherosclerotic lesion. **k**. Adjacent tissue. NC, necrotic core; HR, hemorrhage; C, calcification; CT, cap thinning. Hematoxylin and eosin staining, ×200 magnification

The proportion of macrophages (CD45^+^CD68^+^) among CD45^+^ leukocytes did not differ between the stages of atherosclerotic lesions and was 0.5 (0.2; 0.6)%, but the proportion of macrophages among CD45^+^ leukocytes was significantly higher in AAL samples [6.7 (5.0; 7.8)%] than in EAL samples [3.0 (1.9; 3.6)%; p = 0.003; Fig. 2b]. Additionally, CD45^+^ leukocyte content tended to negatively correlate with the histological type of atherosclerotic lesions (Pearson’s linear correlation coefficient r = −0.57, p = 0.178; Spearman’s correlation coefficient ρ = −0.73, p = 0.064; Fig. 2g).

We also noticed a rank correlation of the number of CD68^+^ macrophages with the numbers of ECs (ρ = 0.58, p = 0.04), contractile VSMCs (ρ = 0.63, p = 0.02), and macrophage-like VSMCs (r = 0.81, p < 0.001; ρ = 0.64, p = 0.02; Fig. 2g).

### Nonimmune cells in EALs and AALs

Here, we quantified two resident cell types of the artery wall: VSMCs and ECs. We used a common marker of monocytic phagocytes—macrosialin (CD68^+^)—to separate contractile VSMCs (CD45^−^αSMA^+^CD68^−^ cells) and intermediate macrophage-like VSMCs (CD45^−^αSMA^+^CD68^+^ cells). ECs were identified as CD45^−^αSMA^−^CD31^+^ cells (Fig. 1a).

The proportion of VSMCs among resident cells of the human coronary artery was 23.5 (12.9; 33.4)%, and the proportion of ECs was 0.6 (0.4; 2.0)%. Contractile VSMCs were the predominant subpopulation of common VSMCs: 90.61 (64.5; 96.2)% in EALs and a slightly lower proportion in AALs [71 (57.1; 90.5)%, p = 0.414]. We did not find any differences in the proportion of either ECs or VSMCs between EALs and AALs (Table 2 and Fig. 2c–f).

Macrophage-like VSMCs were detectable in all samples of coronary atherosclerotic lesions, and the proportion of these cells was 1.9 (0.8; 2.9)% of all cells [7.9 (3.1; 33.2)% in the total VSMC population] in EALs and 3.7 (1.0; 9.1)% of all cells [26.7 (7.8; 41.3)% of all VSMCs] in AAL samples (Table 2).

Even though the number of common VSMCs remained nearly constant from EALs to AALs, a slight shift in the ratios of contractile and macrophage-like subpopulations was observed. Although the proportion of contractile VSMCs tended to be lower in AALs than in EALs, the proportion of macrophage-like VSMCs tended to be higher, albeit statistically insignificantly (p = 0.414 for both comparisons). Additionally, the percentage of macrophage-like VSMCs positively correlated with the histological type of atherosclerotic lesions (r = 0.81, p = 0.029; ρ = 0.87, p = 0.010; Fig. 2g, h).

### Inter- and intraindividual variation of cellular composition of EALs and AALs

Accumulation of VSMCs and ECs was associated with an increased macrophage number in two patients (Patients #2 and #3; Fig. 2i), whereas the other patients did not show this elevation.

We had an opportunity to assess cellular composition in several advanced coronary atherosclerotic plaques (2 to 3 per patient) in four donors (Patients # 1–4; Fig. 2i). AALs manifested high variation of cellular composition within the same artery sample. The highest variation of cell numbers was registered in terms of contractile VSMCs (7-fold on average) and in terms of total VSMCs, macrophages, and ECs (5-fold on average). The proportions of macrophage-like VSMCs were very similar among the plaques within each artery sample (each patient).

### A case of a coronary sirolimus-eluting stent implanted 10 months before death

We investigated the case of in-stent restenosis that occurred 10 months after sirolimus-eluting coronary stenting. The restenosis was the cause of myocardial infarction (Table 1, case ID 4). Mechanisms of rapid neoatherosclerosis are unclear at present, partly due to the lack of information regarding cellular composition of stented arteries. Common mechanisms of restenosis include VSMC proliferation, endothelial dysfunction, and development of inflammation within a short period owing to vascular injury.^34^

We compared the stented coronary artery (including neointima and underlying media) with initial atherosclerotic alterations in the vicinity of the stented region. During the histological analysis of stented segment, we documented a specific sign of neoatherosclerosis: a cholesterol-rich necrotic core with hemorrhages, calcification, and a thin cap (Fig. 2j). Adjacent tissue featured intima thickening and differential proliferation of cell types (Fig. 2k).

Analysis of cell composition of the tissues adjacent to stented and nonstented arteries by flow cytometry did not show any differences (Table 3). By contrast, the total macrophage content and macrophage proportion among leukocytes were significantly higher (5.3- and 5.4-fold, respectively) in the stented artery than in nonstented AAL samples. In particular, leukocytes of the stented plaque contained almost 40% of macrophages, whereas in AALs, this proportion was almost 7% [6.89 (5.0; 7.8)%].

**Table 3.** Cell numbers in atherosclerotic lesions in sirolimus-eluting stented and nonstented coronary arteries

| Cell types | Nonstented arteries |  | Segment adjacent to the stent | Sirolimus-eluting stented artery | P value EAL vs segment adjacent to stent | P value AAL vs sirolimus-eluting stented artery |
| --- | --- | --- | --- | --- | --- | --- |
|  | EAL (n=5) | AAL (n=11) |  |  |  |  |
| Leukocytes (% of singlets) | 15.7 (11.9; 16.1) | 8.1 (3.9; 8.6) | 3.9 | 7.8 | 0.125 | 0.910 |
| Macrophages (% of singlets) | 0.39 (0.28; 0.55) | 0.59 (0.2; 0.66) | 1.1 | 3.7 | 0.125 | 0.004* |
| Macrophages (% of leukocytes) | 3.04 (1.9; 3.6) | 6.89 (5.0; 7.8) | 28.2 | 37.4 | 0.125 | 0.004* |
| VSMCs (% of singlets) | 17.6 (10; 41.5) | 24 (10; 35.5) | 34.5 | 29.9 | 0.250 | 0.250 |
| Contractile VSMCs (% of singlets) | 15.9 (7.3; 39.7) | 15.7 (7.2; 26.8) | 15.1 | 19.3 | 0.625 | 0.652 |
| Contractile VSMCs (% of all VSMCs) | 90.6 (64.5; 96.2) | 71.0 (57.1; 90.5) | 43.7 | 65.5 | 0.125 | 0.359 |
| Macrophage-like VSMCs (% of singlets) | 1.9 (0.8; 2.9) | 3.7 (1; 9.1) | 18.9 | 9.9 | 0.125 | 0.020* |
| Macrophage-like VSMCs (% of all VSMCs) | 7.9 (3.1; 33.2) | 26.7 (7.8; 41.3) | 54.8 | 32.3 | 0.125 | 0.359 |
| ECs (% of singlets) | 0.4 (0.3; 1.4) | 0.6 (0.5; 2.8) | 1.1 | 4.7 | 0.250 | 0.008* |

AAL, advanced atherosclerotic lesion; EAL, early atherosclerotic lesion; ECs: endothelial cells; VSMCs, vascular smooth muscle cells, \*statistically significant.

Furthermore, we noted significantly more macrophage-like VSMCs in the stented plaque (9.9%) than in nonstented AALs [3.7 (1; 9.1)%]. The relative number of macrophage-like VSMCs in the segment adjacent to the stent was also remarkably higher (18.9%) than that in EALs [1.9 (0.8; 2.9)%], without significance.

Finally, EC content of the stented plaque (4.7%) was significantly higher in comparison with nonstented AALs [0.6 (0.5; 2.8)%, p = 0.008; Table 3].

## DISCUSSION

The atherosclerotic plaque is a complex structure composed of at least seven main cell types of various phenotypes and an extracellular matrix and involves cell–cell interactions. It is well known that immune cells play an important role in atherosclerosis.

At first, we compared the numbers of leukocytes (CD45^+^) and macrophages (CD45^+^CD68^+^) between early and advanced human coronary atherosclerotic lesions by flow cytometry. There was a significantly lower relative number of CD45^+^ cells in AALs (8.1%) than in EALs (15.7%).

The depletion of lesional CD45^+^ cells can be driven by cell death, egress, and limited cell recruitment. A study on leukocyte dynamics during atherogenesis in ApoE^−/−^ mice has revealed increased cell death in carotid plaques, corresponding to decreased CD45 staining.^35^ The current theory of progressive atherogenesis includes secondary necrosis, which occurs after lipid loading of macrophages and a loss of their ability to utilize lipids.^36^ Our knowledge about the death of T and B cells, DCs, and other leukocyte subpopulations is much more limited as compared to macrophages.^37^

Another type of cell death is apoptosis, which performs an ambiguous function in atherogenesis, depending on the cell type and stage.^38^ In early studies on human atherosclerosis, plenty of apoptotic immune cells turned out to be located in the shoulders and necrotic core of the plaque.^39^ Macrophage apoptosis promotes the regression of early atherosclerotic lesions but also contributes to necrotic-core enlargement and vulnerability of the advanced atherosclerotic plaque.^40^

In our study, cell egress from plaques cannot be ruled out as a potential mechanism of CD45^+^ cell loss in the atherosclerotic lesion. Reduced leukocyte influx and the enhanced egress initiate inflammation resolution. Macrophages, especially resident macrophages, rather poorly emigrate from tissues. Among them, DCs are best known for clearing from organs by emigrating to draining lymph nodes upon activation.^41^ On the contrary, it has been demonstrated that reverse transendothelial migration of neutrophils is promoted by macrophage interaction via redox-regulated Src family kinase signaling in response to reactive oxygen species.^42^ Leukocyte egress can have a beneficial effect on atherosclerosis resolution but reversely migrating neutrophils can promote systemic inflammation and tissue damage.^43^

In our study, CD45^+^CD68^+^ cell relative numbers among all cells remained unchanged from EALs to AALs (0.39–0.59% of all cells and 3–7% of leukocytes), but the proportion of these cells among leukocytes was significantly higher in the advanced lesions (7%) than in early ones (3%, Fig. 3a). We propose that the immune cell disproportion in atherosclerotic plaques predominantly occurred because of non-CD68^+^ leukocytes, probably, T lymphocytes (as the largest leukocyte population in human AALs^27^) or other cells.

**Figure 3.**
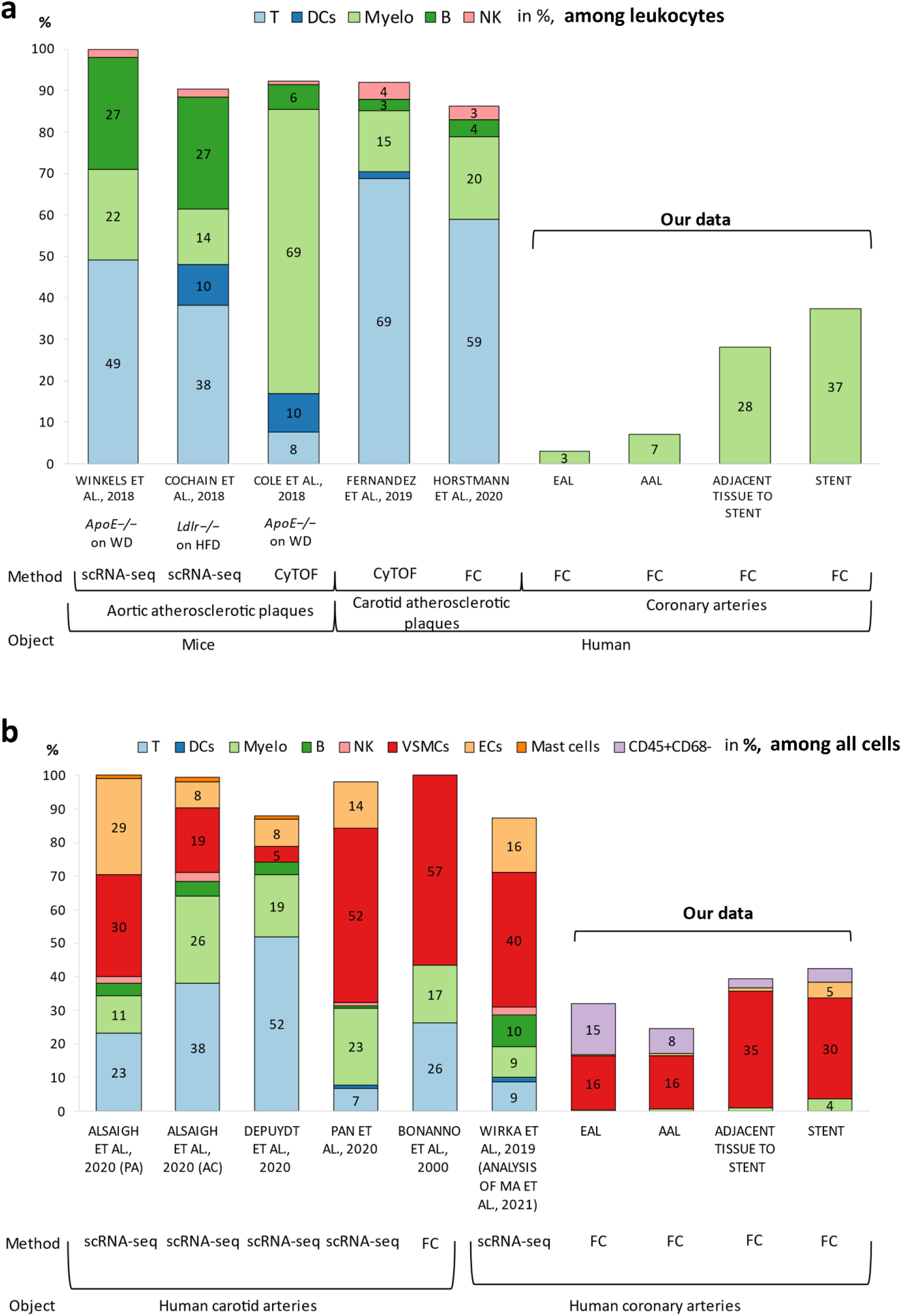
**a**. Immune-cell composition of murine aortic^17,44,45^ and human^25,26^ atherosclerotic lesions. **b**. Cellular composition of human carotid^23,24,27,28^ and coronary atherosclerotic lesions^11,46^. Alsaigh et al. studied a proximal tissue adjacent to a plaque (PA) and a carotid atherosclerotic plaque (AC). EAL: early atherosclerotic lesion, AAL: advanced atherosclerotic lesion. FC: flow cytometry, CyTOF: mass cytometry, scRNA-seq: single-cell RNA sequencing; T: T cells, DCs: dendritic cells, Myelo: myeloid cells, B: B cells, NK: natural killers, WD: Western diet, and HFD: high-fat diet.

Studies on the immune-cell subsets and cellular composition of aortic atherosclerotic lesions in mouse models and human carotid and coronary arteries by cytometry or cell-sorting methods (flow cytometry, CyTOF, and scRNA-seq) have yielded discrepant results due to differences in study design, methodological approaches, and biomaterial sources^11,16,17,23–28,35,44–46^ (Fig. 3a and b). There are mostly macrophages and myeloid cells (14–69%) plus some T cells (8–49%) and B cells (6–27%) among leukocytes in murine aortic atherosclerotic lesions; these variations are a consequence of substantial differences among mouse strains, in tissue digestion and cell isolation protocols, and in leukocyte marker panels used in different studies^17,44,45^ (Fig. 3a). In our study, human early and advanced coronary lesions contained 3–7% of macrophages in the leukocyte population (Fig. 3a). These results contradict previously published percentages of macrophages and myeloid cells in carotid advanced atherosclerotic plaques (15–20%^25,26^; Fig. 3a). Different arteries, cell isolation protocols, leukocyte markers, and methods were employed by us and other authors,^25,26,29^ making a direct comparison between the studies challenging.

Unfortunately, most research articles on human atherosclerotic lesions represent single–time point analyses, mainly of advanced stages (Fig. 3a and b). Nonetheless, the examination of carotid arteries by Alsaigh T. et al. (2020)^24^ revealed increased percentages of myeloid cells among all cells in a carotid atherosclerotic plaque (26%) in comparison to a proximal adjacent tissue (11%). This pathological upregulation of myeloid cells in carotid arteries is consistent with our findings about the progression from EALs to AALs in coronary arteries (3% to 7% of myeloid cells in the leukocyte population; Fig. 3a).

Additionally, the most common specimens for studying human atherosclerosis are carotid endarterectomy samples, which contain intima with a plaque and little media. It should be noted that in our study, T cells and myeloid cells were the main subpopulations of leukocytes, at 59–69% and 15–20%, and of all cells in carotid plaques, at 7–52% and 11– 26%, respectively (Fig. 3a and b). The proportion of B cells was 3–4% among leukocytes in carotid atherosclerotic plaques (Fig. 3a). Moreover, VSMCs made up 5–57% of all cells in carotid plaques (Fig. 3b).

In this study, we analyzed whole coronary arteries, which are different from carotid arteries in their morphology and temporal and spatial cellular patterns.^47^ Compared with carotid plaques, the coronary arteries of patients with atherosclerosis turned out to be enriched in VSMCs, which in this tissue constitute 16–35% and 40% of all cells according to our data and other studies,^11^ respectively (Fig. 3b). The proportions of T, B, and myeloid cells were nearly equal in coronary arteries (approximately 10% for each cell type).^46^

Media and adventitia of atherosclerotic plaques contain most of VSMCs and B cells; therefore, in a carotid lesion that lacks these tissues, counts of some leukocytes are overestimated in comparison with a whole artery. Winkels et al. (2018) found predominant accumulation of myeloid cells and T cells in microdissected mouse aortic plaques and human carotid plaques while B cells predominated in the adventitia and proved to be only a minor subpopulation in plaques.^44^ In the present work, we isolated cells from whole coronary arteries encompassing intima, media, and adventitia; hence, we registered higher prevalence of VSMCs and a smaller proportion of macrophages as compared with carotid arteries.

Disproportionate amounts of immune cells in arteries depend on the vascular bed.^48^ Here, we identified differences in leukocyte composition and in the proportion of macrophages not only between EALs and AALs but also among different individuals and among AALs in some patients.

It is reported that contractile αSMA^+^ cells can gain a capacity for lipid loading via so-called transdifferentiation.^36^ During the phenotypic switching, they acquire macrophage-like properties, including CD68 expression, as confirmed in a murine atherosclerosis model.^49^ Coexpression of αSMA (also known as ACTA2) and CD68 in smooth muscle cells is observed in lineage-tracking murine models, whereas 16% of CD68^+^ cells may have nonmyeloid origin.^50^

In human advanced coronary plaques, Allahverdian and coworkers (2014) detected costaining of αSMA and lipids (oil red O staining) in 50% ± 7% of all foam cells. Furthermore, they noticed that 40% ± 6% of all CD68^+^ cells also expressed αSMA.^13^ On the other hand, this number may include both myeloid and VSMC origin cells owing to a lack of CD45 staining.

In this work, we revealed that the relative number of CD45^−^αSMA^+^CD68^+^ cells was 2– 4% and 8–27% of all αSMA^+^ cells in early and late coronary atherosclerotic lesions, respectively, demonstrating that this cell phenotype is present at early stages of human coronary lesions. Moreover, the number of macrophage-like VSMCs significantly correlated with the number of CD45^+^CD68^+^ macrophages (r = 0.81; ρ = 0.64; p < 0.05) and the histological type of an atherosclerotic lesion (r = 0.81; ρ = 0.87; p < 0.05). These results are in agreement with the existing knowledge about macrophage–VSMC interactions and cell phenotypic switching.^51^ In another study, cocultivation of VSMCs and macrophages revealed a proinflammatory influence of the latter, stimulation of VSMC proliferation, and acquisition of a procalcific phenotype by VSMCs.^52^ Nonetheless, the specific stimuli causing VSMCs to acquire the macrophage-like phenotype remain unclear.

We demonstrated notable differences in the numbers of some cell types between nonstented and sirolimus-eluting stented segments of human coronary atherosclerotic plaques, especially in the proportion of ECs (0.6% vs 4.7% of singlets), macrophages (0.59% vs 3.7% of singlets and 6.89% vs. 37.4% of leukocytes), and macrophage-like VSMCs (3.7% vs. 9.9% of all cells), while the proportions of contractile VSMCs were similar (Table 3 and Fig. 3). Additionally, we did not find statistically significant differences in cellular composition between early plaques and the arterial tissue adjacent to the stent.

Our results are consistent with data from human autopsy studies on drug-eluting stents in coronary arteries. Restenotic samples offer evidence of hypercellularity or ongoing cellular proliferation.^53,54^ It is reported that the main driver of in-stent restenosis is VSMC dedifferentiation and proliferation accompanying macrophage infiltration and neointimal angiogenesis.^55^ By contrast, Schwartz et al. (1996) hypothesized that restenotic arteries are characterized by cellular proliferation only at early stages; consequently, lesions derived from human coronary arteries are not characterized by cellular proliferation if sampled at later stages.^56^ Therefore, further studies with larger sample size are needed to test this hypothesis.

One of the limitations of this study is small sample size. Nonetheless, some of our results are statistically significant even with 16 specimens of coronary artery plaques. Additionally, the enzymatic tissue digestion for the isolation of cells from an atherosclerotic lesion may induce cell death or other enzymatic-extraction–related artifacts that may affect flow-cytometric results in a batch-dependent manner. Besides, the cellular composition of coronary arteries in this work was not verified by other methods.

Although all the analyzed patients were clinically symptomatic (had acute coronary syndrome), AALs in this study were stable according to histological analysis, except for two samples in one case of restenotic coronary lesions. To confirm the significance and generality of our findings, further research would be warranted with larger numbers of atherosclerotic lesions that differ in histological vulnerability and vascular beds among patients’ subgroups (e.g., clinically symptomatic/asymptomatic and diabetic/nondiabetic).

Here, we for the first time investigated ECs (CD45^−^αSMA^−^CD31^+^), leukocytes (CD45^+^) and separately macrophages (CD45^+^CD68^+^) and contractile (CD45^−^αSMA^+^CD68^−^) and macrophage-like (CD45^−^αSMA^+^CD68^+^) VSMCs in EALs and AALs of human coronary arteries by flow cytometry. We documented differences in CD45^+^ leukocyte and CD45^+^CD68^+^ macrophage numbers between EALs and AALs. The proportion of CD45^+^ leukocytes diminished, while the subpopulation of CD45^+^CD68^+^ macrophages expanded in advanced coronary atherosclerotic plaques compared with the early lesions. We detected αSMA^+^CD68^+^ VSMCs in EALs. There was also a strong positive correlation of the αSMA^+^CD68^+^ VSMC number with the CD45^+^CD68^+^ macrophage number and with the histological type of atherosclerotic lesions. The restenotic coronary lesion was found to contain large numbers of ECs, macrophages, and αSMA^+^CD68^+^ VSMCs. In conclusion, we would like to emphasize the pivotal role of complex changes in the numbers of adaptive-immunity cells and macrophage-like VSMCs during human coronary atherosclerosis.

## Data Availability

All the data have been presented in the manuscript

## Abbreviations

VSMCs: Vascular smooth muscle cells
ECS: Endothelial cells
DCs: Dendritic cells
ScRNA-Seq: Single-cell RNA sequencing
EAL: Early atherosclerotic lesion
AAL: Advanced atherosclerotic lesion

## Acknowledgments

The authors thank a pathologist of Siberian State Medical University, Nadezhda Krahmal, for her help with the biomaterial collection. The English language was corrected and certified by shevchuk-editing.com.

